# A Fundamental Inconsistency in the SIR Model Structure and Proposed Remedies

**DOI:** 10.1101/2020.04.26.20080960

**Authors:** Michael Nikolaou

## Abstract

The susceptible-infectious-removed (SIR) compartmental model structure and its variants are a fundamental modeling tool in epidemiology. As typically used, however, this tool may introduce an inconsistency by assuming that the rate of depletion of a compartment is proportional to the content of that compartment. As mentioned in the seminal SIR work of Kermack and McKendrick, this is an assumption of mathematical convenience rather than realism. As such, it leads to underprediction of the infectious compartment peaks by a factor of about two, a problem of particular importance when dealing with availability of resources during an epidemic. To remedy this problem, we develop the dSIR model structure, comprising a single delay differential equation and associated delay algebraic equations. We show that SIR and dSIR fully agree in assessing stability and long-term values of a population through an epidemic, but differ considerably in the exponential rates of ascent and descent as well as peak values during the epidemic. The novel Padé-SIR structure is also introduced as a approximation of dSIR by ordinary differential equations. We rigorously analyze the properties of these models and present a number of illustrative simulations, particularly in view of the recent coronavirus epidemic. Suggestions for further study are made.

## 1. Introduction

In their landmark 1927 publication *Contribution to the Mathematical Theory of Epidemics*,^1,2^ Kermack and McKendrick developed a general, if elaborate model structure to capture the dynamics of a fixed-size population comprising compartments of individuals susceptible (S) to a spreading infection, infectious (I), and removed (R) from the preceding two compartments by recovery or death. Propagation of an individual from S to I to R underlies the basic context of the exercise. In a modeling tour-de-force, the authors eventually present, in equations (11) through (15) of their paper (*ibid*.), the general structure of the elaborate mathematical model they derive. They proceed to examine the implications of their model for special cases, and finally present a very special case resulting in a set of three relatively simple ordinary differential equations (ODEs, equations (29) *ibid*.), which were destined to form the basis for a genre of mathematical models in epidemiology, the celebrated SIR model and its many variants.^3^ The three ODEs are

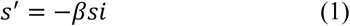

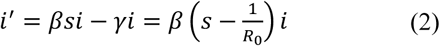

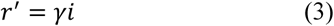

where prime denotes time derivative.^2,4,5^ Consistent with the importance of the SIR ODEs, the *basic reproductive ratio*,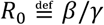, is widely considered “one of the most critical epidemiological parameters”^2^ and has even become a household name in the recent coronavirus epidemic.^6^

In the sentence right before they present their SIR model in equation (29), (*ibid*., cf. the above eqns. (1)-(3)) Kermack and McKendrick explain that this is a

> *special case in which ϕ and ψ are constants κ and l respectively*

with (*κ, l*) refering to (*β, γ*) of eqns. (1)-(3), respectively.

The assumption about constant *ϕ* is plausible, as it refers to the rate of spread of the epidemic (cf. eqn. (1)). While that parameter might change over time as a result of measures taken to curb an epidemic, such a change could easily be reflected in the SIR model by a time-varying *ϕ* (cf. *β* in eqns. (1) and (2)).

The assumption about constant *ψ*, however, as fruitful as it may have proved, is chosen for mathematical convenience rather than for intent to describe the system as realistically as possible. Indeed, a clear definition of *ψ* is provided by the authors as follows (p. 703, *ibid*.):

> *If ψ*_θ_ *denotes the rate of removal, then the number who are removed from each θ group at the end of the interval t is ψ_θ_ν_t,θ_*,

where (p. 702, *ibid*.)

> *ν_t,_*_θ_ *shall denote the number of individuals in unit area at the time t who have been infected for θ intervals*.

However, the rate of removal depends more on the duration over which individuals have remained infected and less on the size of that group. In the conceptually simple case where all individuals recover or die at a single number of days, *D*, after their infection, the rate of removal is 1 or 0, depending on whether *θ* is greater than *D* or not (Figure 1).

**Figure 1.**
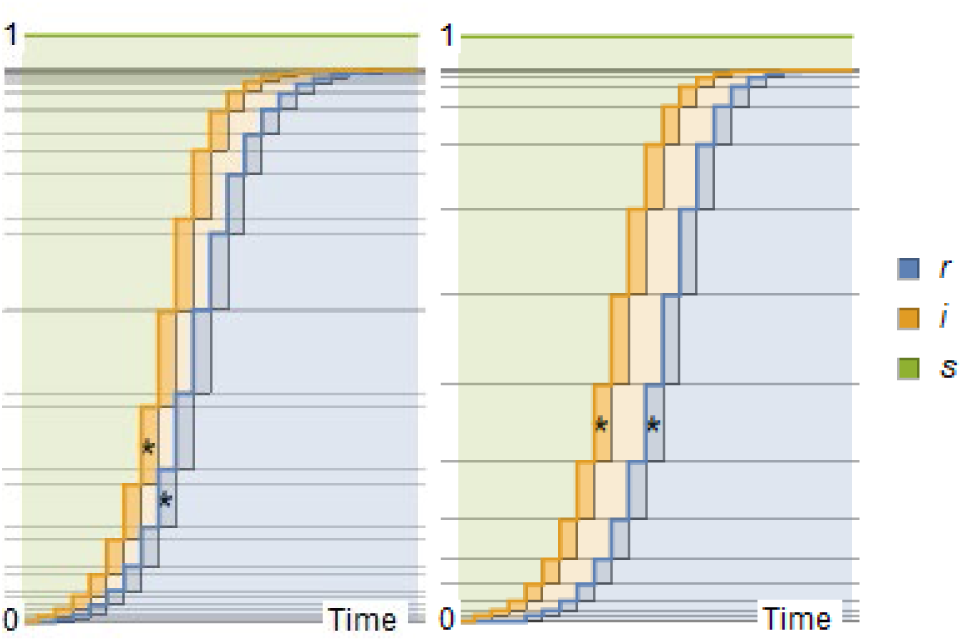
*Stacked profiles of* {*s*, *i, r*} *over discrete time in a constant-size population over time. Left: At each time step, r increases in proportion to the previous i. Right: At each time step, r increases by the decrease in s at a fixed number of steps before. Stars indicate correspondence in the i to r transition*.

Of course, in reality *D* will likely follow a distribution (Figure 2) rather than being a single number. However, in that case as well, the removal rate will depend on comparison between *θ* and the distribution of *D*, rather than on the size of the group remaining infectious for time *θ*.

Starting with the assumption that individuals leave the infectious group at time *D* after infection, we develop in this paper a corresponding mathematical model structure, named delay SIR (dSIR), in the form of a single delay differential equation (DDE) for *s*, and two associated delay algebraic equations, for *i* and *r* in terms of *s*.

In the rest of the paper we first introduce the dSIR model structure in section 2, and provide an intuitive exposition of its basic properties in section 3, where we also introduce the Padé SIR model structure as an ODE approximation of dSIR. Rigorous analysis follows in section 4. In that section we explain that certain SIR and dSIR properties are exactly similar (e.g. herd immunity, total number of infected), while others are quite different (e.g. exponential rates, peak of the infectious group). Extension to models with additional compartments beyond S, I, and R is discussed in section 5, with presentation of the dSPIR model. Finally, the significance of this work and future extensions are discussed.

**Figure 2.**
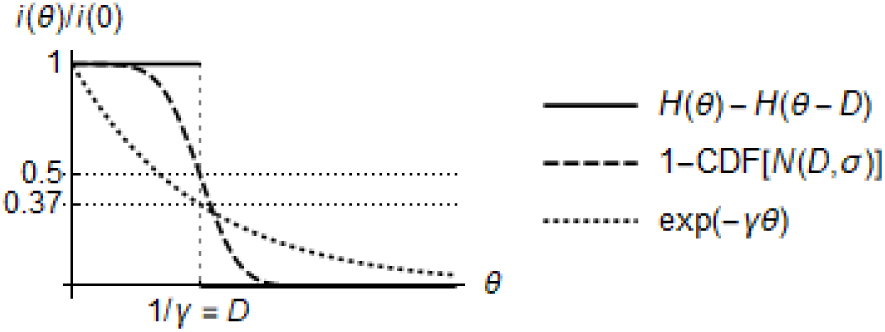
*Reduction of the infectious fraction i from i*(0) *to* 0 (*a*) *in a single front in terms of the Heaviside step function H with D =* 1*/γ*, (b) *in a diffused front in terms of the cumulative density function* (*CDF*) *of the normal distribution N*(*D, σ*)*, and* (*c*) *at rate* −*γi, yielding exponential decline, eqn*. (2).

## 2. The dSIR model structure

To explain the derivation of the dSIR model structure, we will rely on the detailed version of Figure 2 shown in Figure 3. The schematic shows the evolution of *s_k_, i_k_, r_k_* over discrete time steps *k* = 0,1,2,… of length *δt* each.

The thick-green bordered rectangle in Figure 3 suggests by visual inspection that

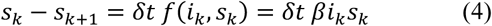

where *f*(*r_k_, s_k_*) is the rate at which the infection spreads, for example in proportion to the product *i_k_s_k_*, as shown above.

Assuming that each new part of the infectious fraction, *i_k_*, gained at a given time step, *k*, moves to the removed fraction, *r*, in *n* time steps, the thick-red bordered rectangle in Figure 3 suggests that

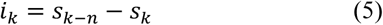

Eqns. (4) and (5) yield

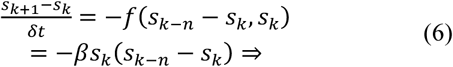

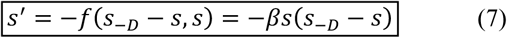

where 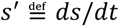 and 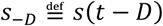.

Eqn. (7) is a single nonlinear DDE that involves only *s* and is decoupled from equations for *i* and *r*. As such, it captures the entire dynamics of the dSIR system.

The remaining two fractions of the population, *i* and *r*, can simply be inferred by algebraic equations, as eqn. (5) implies

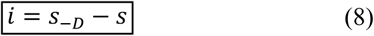

and *i + s + r* = 1 implies

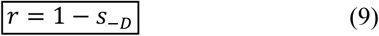

**Figure 3.**
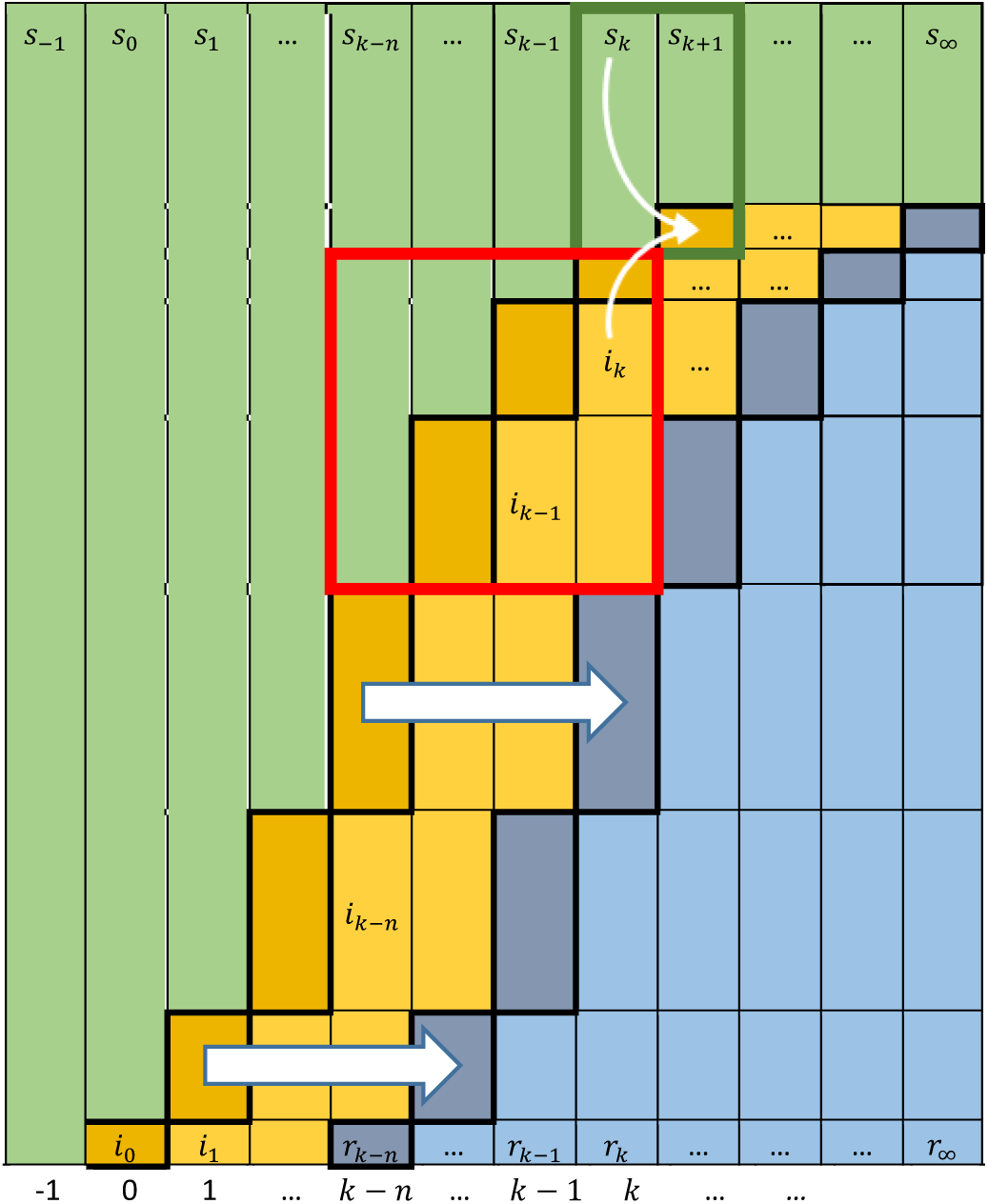
*Schematic of evolving susceptible* (*green*)*, infectious* (*orange*)*, and removed* (*blue*) *fractions of a fixed-size population after an initial infection, i*_0_*. Each new part of the infectious fraction* (*thick-black bordered orange rectangles*) *moves to the removed fraction* (*thick-black bordered blue rectangles*) *in n time steps. The population eventually reaches a steady state at s_∞_, r_∞_ =* 1 − *s_∞_, and i_∞_ =* 0.

To put the schematic in Figure 3 in context, observe that summation of eqns. (1)-(3) and discretization yields

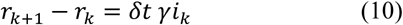

This suggests that, according to the SIR model, each thick-black bordered green rectangle in Figure 3 would have to be a *constant* fraction *δt γ* of the immediately previous orange column. However, with each new part of the infectious fraction moving to the removed fraction after *n* steps (thick-black bordered orange rectangles moving to thick-black bordered blue rectangles) this cannot be true as an emerging fact, by the simple observation of the shapes of *r* and *r* + *i*, which follow a convex to concave pattern after an inflection point. Therefore, the assumption of constant *ψ*, equivalent to constant *γ* in eqn. (10), is not compatible with the assumption of time to transition from infectious to removed being independent of the infectious fraction size. This will be further illustrated with simulations in the next section.

Incidentally, eqns. (1)-(3) of the SIR model can also be decoupled to a single if non-intuitive nonlinear ODE in terms of *s*, solved in a form implicit in time, *t*.^7^

## 3. From SIR to dSIR and beyond: Visualization

Before any theoretical properties of the dSIR model structure are analyzed, a simple visual comparison between SIR and dSIR is presented. Unless specifically stated otherwise, all numerical simulations have been conducted with^8,9^

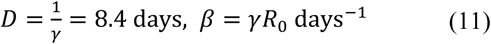

Figure 4 is the standard {*s*, *i, r*} plot, for SIR and dSIR. Note the faster dynamics of dSIR compared to SIR, the approximately twice as high peak of *i* for dSIR compared to SIR, as well as the asymmetric profile of *i* for SIR, compared to the symmetric dSIR profile of *i*. (Details in section 4.)

**Figure 4.**
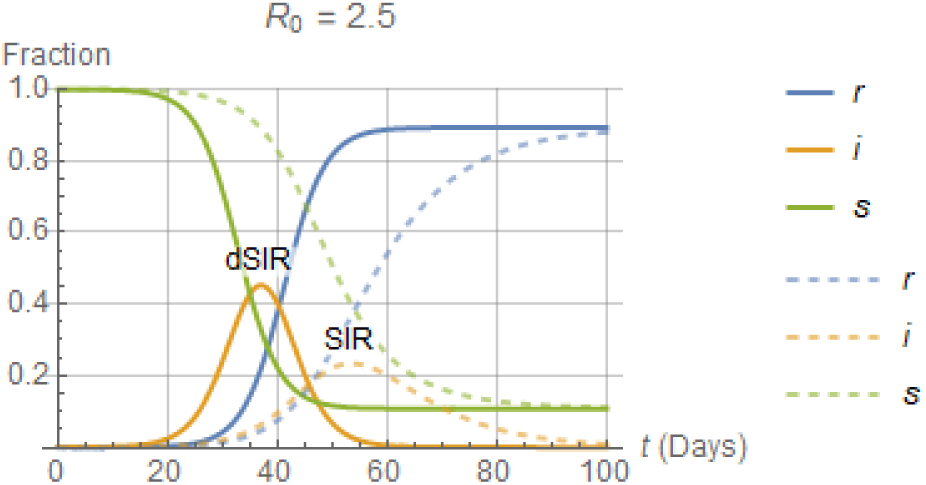
*Profiles of s, i, r of a population through an epidemic according to the dSIR and SIR models, eqns*. (7)-(9) *and* (1)-(3) *respectively. The correspondence γ =* 1/*D was used*.

To further illustrate the dSIR/SIR relationship, Figure 5 and Figure 6 show the continuous-time counterparts of Figure 3 for the SIR and dSIR models, respectively. In this stacked representation of {*s*, *i, r*} over time, it is clear that the SIR model corresponds to a time-varying infectious period for each individual. In fact, the value of *γ* calculated according to eqn. (3) as

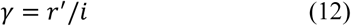

with *r′* and *i* produced by the dSIR model, is time varying, as shown in Figure 7. This discrepancy suggests that the standard interpretation of 1*/γ* as

> *the average infectious period* … *estimated relatively precisely from epidemiological data*^2^

is increasingly inaccurate as *R*_0_ increases above 1. What is estimated from epidemiological data is the delay *D*, rather than *γ*, and if 1/*D* is used as an estimate of *γ*, as is typically done, the SIR model response will be too slow, with a peak value for *i* lower than its dSIR counterpart by about half. Depending on the *R*_0_ value considered, this may significantly affect the SIR model value for “Flattening the Curve”, an aim that has become a household name in the latest coronavirus epidemic.^10^

**Figure 5.**
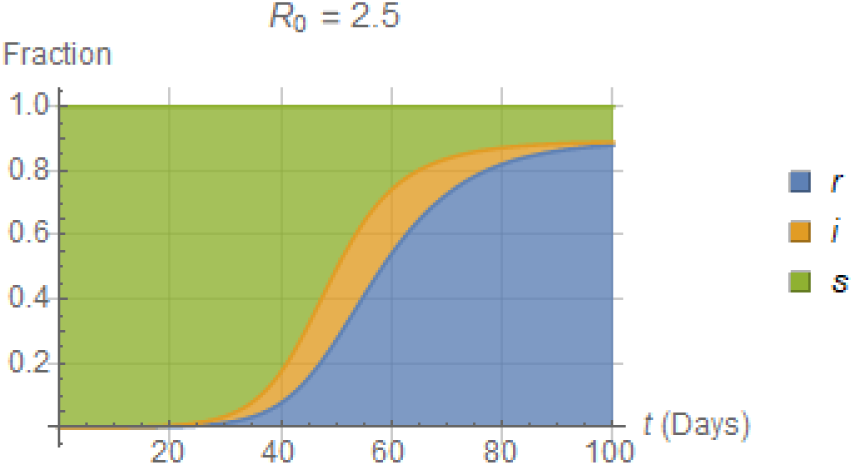
*Stacked fractions* {*s*, *i, r*} *of a population through an epidemic according to the* SIR *model, eqns*. (1)-(3)*. Note that the duration from entrance to the infectious group until removal from it* (*horizontal slices in orange area*) *is variable*.

**Figure 6.**
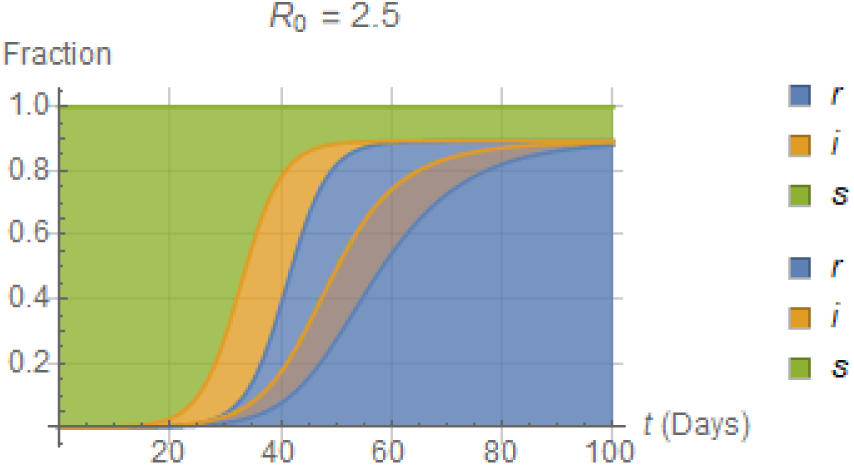
*Stacked fractions* {*s*, *i, r*} *of a population through an epidemic according to the* dSIR *model, eqns*. (7)-(9)*. Note that D remains constant. Figure* 5 *is also superimposed, for comparison. The correspondence γ =* 1/*D was used*.

**Figure 7.**
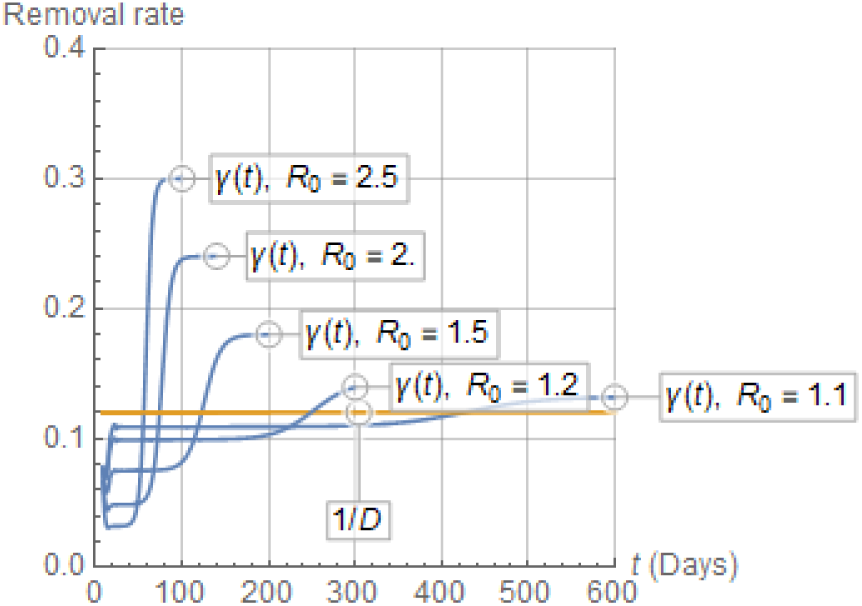
*Comparison between* 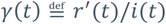 *and the corresponding* 1/*D =* 0.12 *day^−^*^1^ *for various R*_0_ *in numerical integration of the* dSIR *eqns*. (7)-(9). *Note that all lines start at t = D, as r′*(*t < D*) *=* 0*. Note also small spurious deviations from constant shortly after t = D, due to numerical approximation of i*(*t ≤* 0), *by the continuous function* (1 − *∊*) *exp*(*λt*)*, t ≤* 0 *with λ »* 1.

### 3.1 The Padé SIR model structure

As pointed out in the preceding sections, interpreting the value of *γ* in the SIR model as the average infectious period is not accurate and produces misleading results. It turns out (Appendix A) that the following simple remedy can be used to retain the ODE structure of the standard SIR model, while better approximating the DDE dynamics of the more realistic dSIR model structure: The SIR equations for {*i′, s*′}, eqns. (2) and (3), can be replaced by the equally simple ODEs

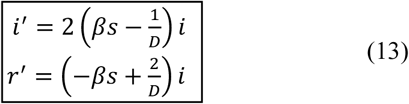

or by the more accurate set of ODEs

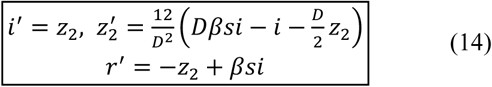

with *z*_2_(0) = 0.

Figure 8 confirms that the *i* trajectories for the dSIR and the modified SIR model structures – eqns. (7)-(9) and (1)-(3) with eqn. (2) replaced by (13) or (14), respectively – are close to one another, both in terms of the time to peak and the value of the peak. As already mentioned, these properties are important when such models are used to anticipate hospitalization needs for the infected during an epidemic.

**Figure 8.**
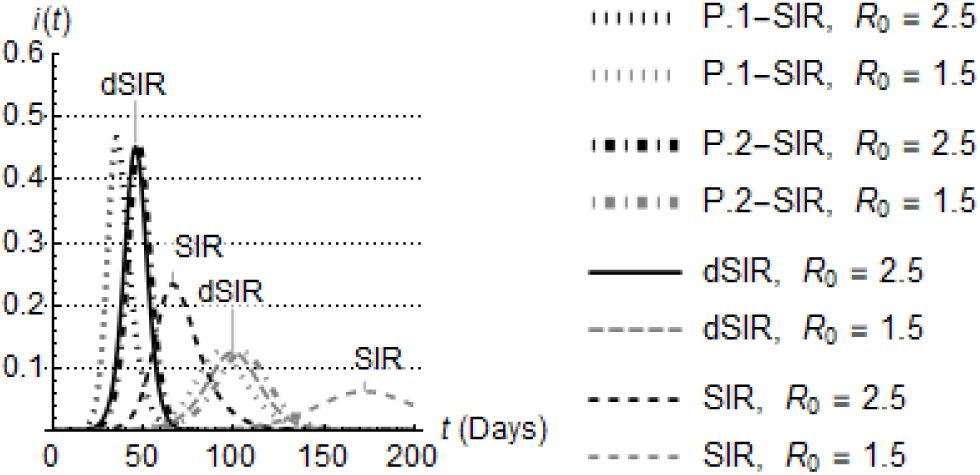
*Comparison of the i profiles for the first-order* Padé SIR, *second-order* Padé SIR, dSIR, *and* SIR *models. Note the improve approximation of* dSIR *by the second-order* Padé SIR, *compared to first-order* Padé SIR, *as anticipated by eqns*. (13) *or* (14)*, respectively*.

Note (Appendix A) that the essence of eqns. (13) and (14) is in approximating the pulse profile *H*(*θ*) − *H*(*θ* − *D*) in Figure 2 by a transfer function approximation based on first-or second-order *Padé-approximants* (in the Laplace domain)

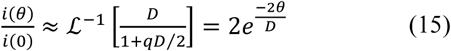

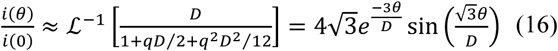

(rather than by the decaying exponential of Figure 2) as shown in Figure 9. Padé-approximation has long been a popular approach for approximating transcendental transfer functions by polynomial rational fractions in process control.^11^

**Figure 9.**
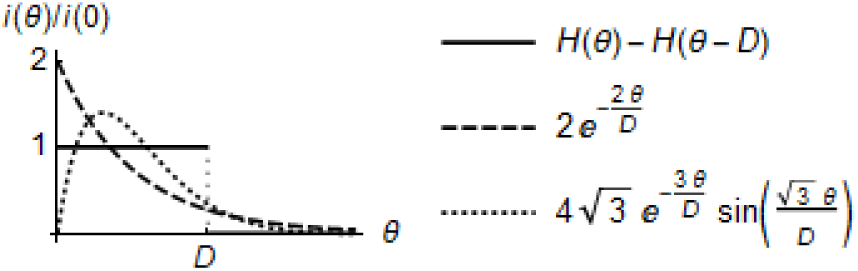
*Reduction of the infectious fraction i from i*(0) *to* 0 (*a*) *in a single front in terms of the Heaviside step function H with D =* 1*/γ*, (b) *following a first-order Padé approximation, and* (c) *following a second-order Padé approximation*. (*cf. Figure* 2).

Because of the critical role of Padé approximation in deriving eqn. (14) for the ODEs of the modified SIR model to approximate the dSIR model, we will use the term Padé SIR to denote the modified SIR model structure.

Finally, it should be noted that eqn. (13) may seem counter-intuitive, as it appears to suggest that the generation and depletion rates of *i* are 2*βsi* and −2*i/D*, respectively. However, eqn. (13) rather suggests that while the susceptible depletion rate remains −*βsi*, the infectious depletion rate appears as (*βs* − 2*/D*)*i* (which is *s*-dependent) rather than −*i/D* (which is not) due simply to replacement of the delay term *e*^−^*^qD^* in the Laplace domain by its Padé approximant. Similarly, the rate of increase for *r* in eqn. (13) is *s*-dependent, rather than not.

## 4. Dynamics of the dSIR and Padé SIR models

Standard theory of DDEs (e.g. equation (9.42) in Ch. 4 and equation (1.2) in Ch. 5 of Kuang,^12^ or Gopalsamy^13^) can be applied to establish rigorous properties for the dSIR model, such as global stability, convergence to a final steady state, and others. A complete analysis is beyond the scope of this paper. However, some important theoretical properties of the dSIR model structure of practical interest are discussed next, particularly in comparison to their SIR counterparts.

The analysis of the Padé SIR model follows standard ODE analysis and is presented more briefly, except when it has important implications for either theoretical or practical issues.

### 4.1 Stability at equilibrium and epidemic outbreak

Eqn. (7) can be used to show (Appendix B) that an equilibrium point 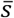 is stable and an epidemic outbreak does *not* occur iff

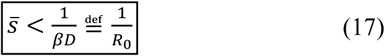

Note that the stability upper bound 1*/*(*βD*) for the dSIR model in the above eqn. (17) coincides with the well known bound γ*/β =* 1*/R*_0_ dictated by the SIR model under the widely used correspondence

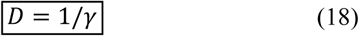

The same stability bound can be derived for the Padé SIR model structure using standard ODE analysis based on linearization around an equilibrium point.

### 4.2 *Final values of* {*s, i, r*}

Accepting for now without proof the global stability of eqn. (7) and existence of (*s_∞_*, *i_∞_* = 0, *r_∞_*} for initial values *i*_0_ *≈ ∊≈* 0, *s*_0_ = 1 − *∊≈* 1, one can show (Appendix C) that

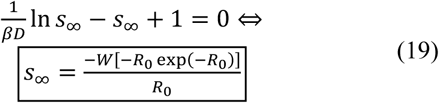

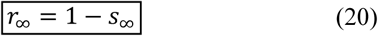

where 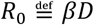 and *W* is the *Lambert function*^14,15^ of order 0.

The standard plot for *r_∞_*, equal to the total fraction of infected by the end of an epidemic,^2^ is shown in Figure 10 for completeness. Note that the plot is valid for both SIR and dSIR models, with *R*_0_ *= β/γ* and *R*_0_ *=* 1/*D*, respectively.

**Figure 10.**
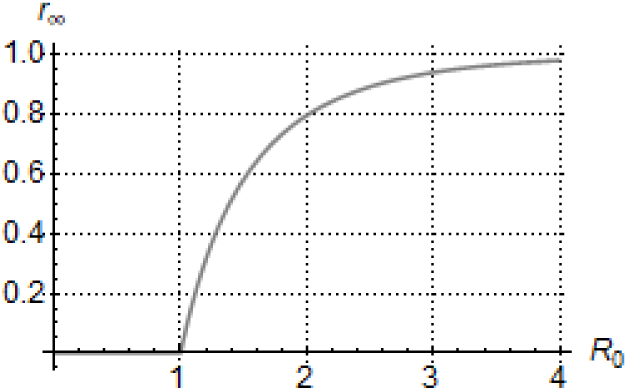
*Total fraction of a population infected by the end of an epidemic, r_∞_, as a function of the basic reproductive ratio R*_0_ *= β/γ =* 1*/D according to eqn*. (20)*, similar for both the* SIR *and* dSIR *model structures. For R*_0_ <1 *the epidemic is contained*.

Interestingly, while use of the Lambert function to solve problems such as the above was pointed out as early as 1996,^14^ it may have escaped the attention of most literature in this field.^2^ The Lambert function in its various forms will turn up in a number of results below.

It should also be noted that eqns. (19) and (20) are the same for the SIR and Padé SIR model (Appendix C) under the correspondence between *D* and *y* in eqn. (18).

### 4.3 Exponential rates of ascent

For the initial part of a spreading epidemic, starting from a perturbation of the steady state 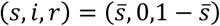 as

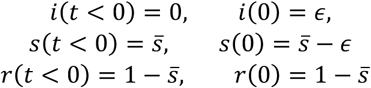

With

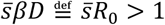

it can be shown (Appendix D) that the infectious fraction initially grows approximately at an exponential rate as

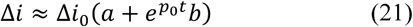

where 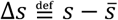 and the constants *a, b* are in terms of 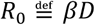 (Appendix D) with

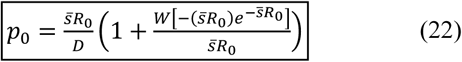

as shown in Figure 11.

Similarly, the initial exponential rate of the Padé SIR model is given by

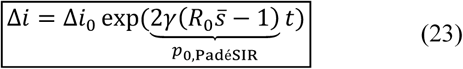

where *R*_0_ *= β/γ*, and for the standard SIR model, the exponential rate is

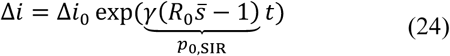

where *R*_0_ *= β/γ*, shown in Figure 11 as well.

**Figure 11.**
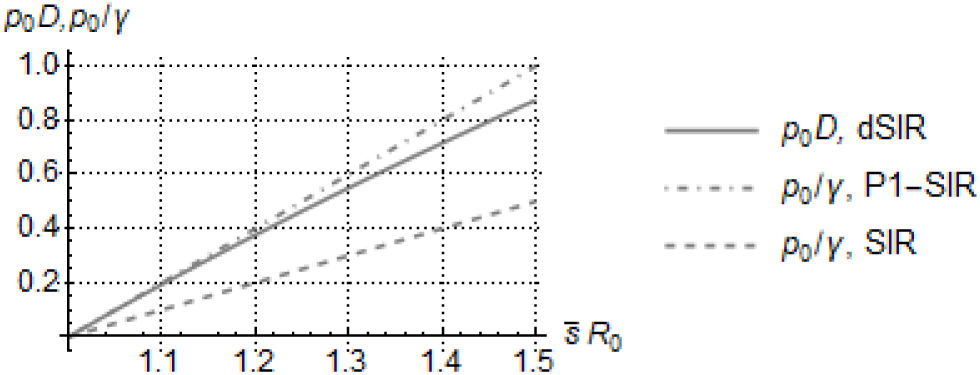
*Dimensionless exponential rates {p*_0_*D, p*_0_*/γ} for the* dSIR, *first-order* Padé SIR, *and* SIR *models, by eqns*. (22), (23)*, and* (24)*, respectively. The first two coincide fairly well as* 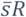 *approaches* 1*, whereas the* SIR *rate remains about half of the other two*.

This has immediate implications for the early rate of rise of the infectious fraction to its peak, *i^*^*, as illustrated in Figure 12.

**Figure 12.**
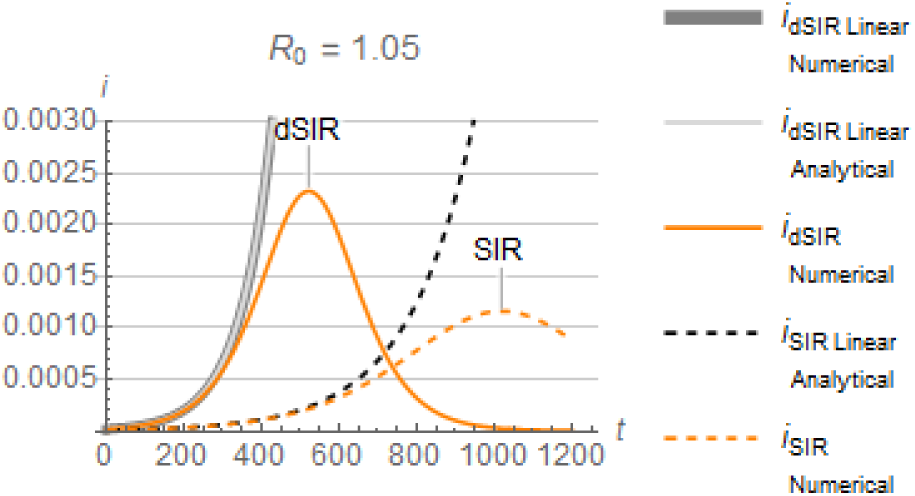
*Exponential rate of increase at the early stages of an epidemic for the* dSIR *and* SIR *model structures*.

Note that while the initial SIR rate is half of the Padé SIR rate and about half of the dSIR rate, all three models eventually reach the exact same steady-state values, as captured by eqns. (19) and (20).

### 4.4 Peak of infectious fraction

While it is not obvious to the author whether the peak *i^*^* can be easily obtained for the dSIR model, a good approximation can be obtained (Appendix E) through the Padé SIR model, as

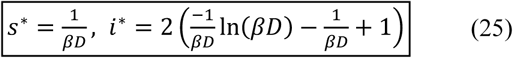

Note that the above *i^*^*, exact for the first-order Padé SIR model and approximate for the second-order Padé SIR and dSIR models, is double the *i^*^* of the standard SIR model, as confirmed in Figure 8. Once more, there are obvious practical implications from this discrepancy.

Note also that in case an upper bound is placed on *i^*^*, to avoid overwhelming hospitalization facilities during an epidemic, eqn. (25) has an explicit solution for the corresponding maximum 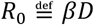 as

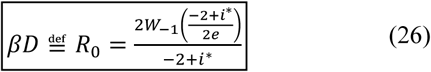

where 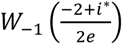 is the Lambert function of order −1.

For comparison, the standard SIR model yields

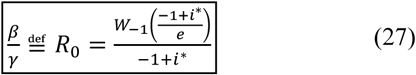

The values of *R*_0_ indicated by eqns. (26) and (27), with corresponding definitions, are shown in Figure 13. It is evident that the Padé SIR model places twice as tight a restriction on *R*_0_ as the standard SIR model, if *i* is not to exceed the *i^*^* value.

**Figure 13.**
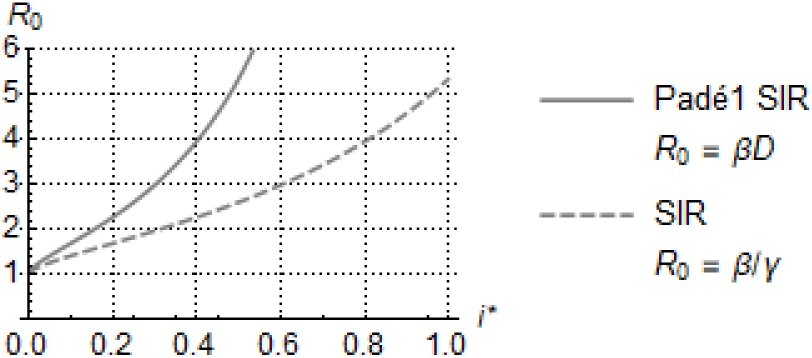
*Maximum value of R*_0_ *indicated by the* Padé SIR *and* SIR *models for i not to exceed i^*^*.

## 5. dSPIR, Padé SPIR, and variants

The dSIR model structure developed in Figure 3 can be easily extended to include additional compartments. In fact, practically all population models of infections developed to date using the concept of exchange between compartments^3^ can be immediately translated (a) from ODEs to DDEs through replacement of compartment drain rates proportional to the drained quantity by drainage of amounts that have resided for a certain time, *D*, in that compartment, or (b) from ODEs to Padé approximations that maintain the ODE structure but are more realistic. We illustrate these ideas on the SPIR model structure, in view of its importance for the recent coronavirus epidemic.^16-18^

A distinct feature of the coronavirus causing COVID-19 is that it enables infection transmission at the pre-symptomatic stage.^19^ Therefore, the SPIR model structure (Figure 14) comprises the usual population fractions {*s*, *i, r*} along with the pre-symptomatic infectious fraction, *p*, with symptomatic infectious being *i*. SPIR differs from the standard SEIR structure^2,4,5,20^ by the way the four compartments interact.

**Figure 14.**
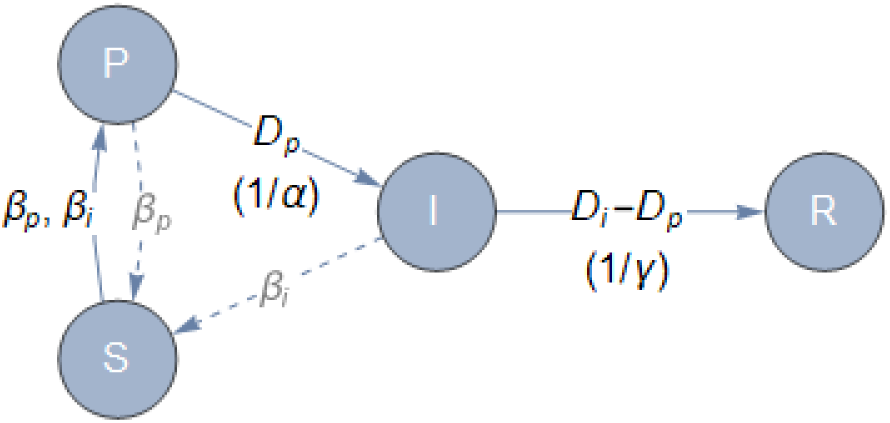
*The* SPIR *and* dSPIR *model structure, with members of the pre-symptomatic infectious compartment*, P, *moving, after time D_p_, to the symptomatic infectious compartment, I, and from there, after time D_i_* − *D_p_, to the removed compartment*, R. *Both* P *and* I *infect the susceptible group*, S, *at rates, β_p_ and β_i_*, *respectively*.

While it is formidable to practically monitor infections on pre-symptomatic infectious individuals, monitoring symptomatic infectious is more reasonable, as symptoms are clear and can be confirmed by testing. Therefore, tighter restrictions can be placed on the I group, in addition to the P group typically following general restrictions placed on the general population to curb the spread of the epidemic, as captured by the spread factors *β_i_* and *β_p_*, respectively, in Figure 14.

The dynamics of the dSPIR structure is shown in Figure 15, which follows the pattern of Figure 3, with the addition of the pre-symptomatic infectious fraction *p*.

Following the same technique as in section 2, we can immediately write the following equations for the dSPIR model structure by visual inspection of Figure 15:

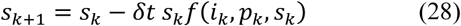

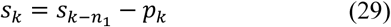

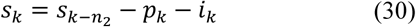

Combining the above equations and letting *δt → ∞* yields the nonlinear DDE

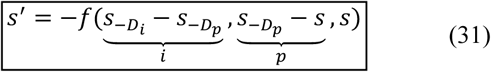

and associated delay algebraic equations

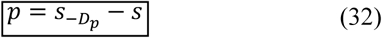

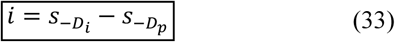

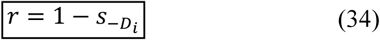

where *D_p_* and *D_i_* − *D_p_* are the durations of an individual’s staying in the P and I compartment, respectively, and typically

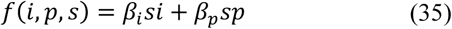

**Figure 15.**
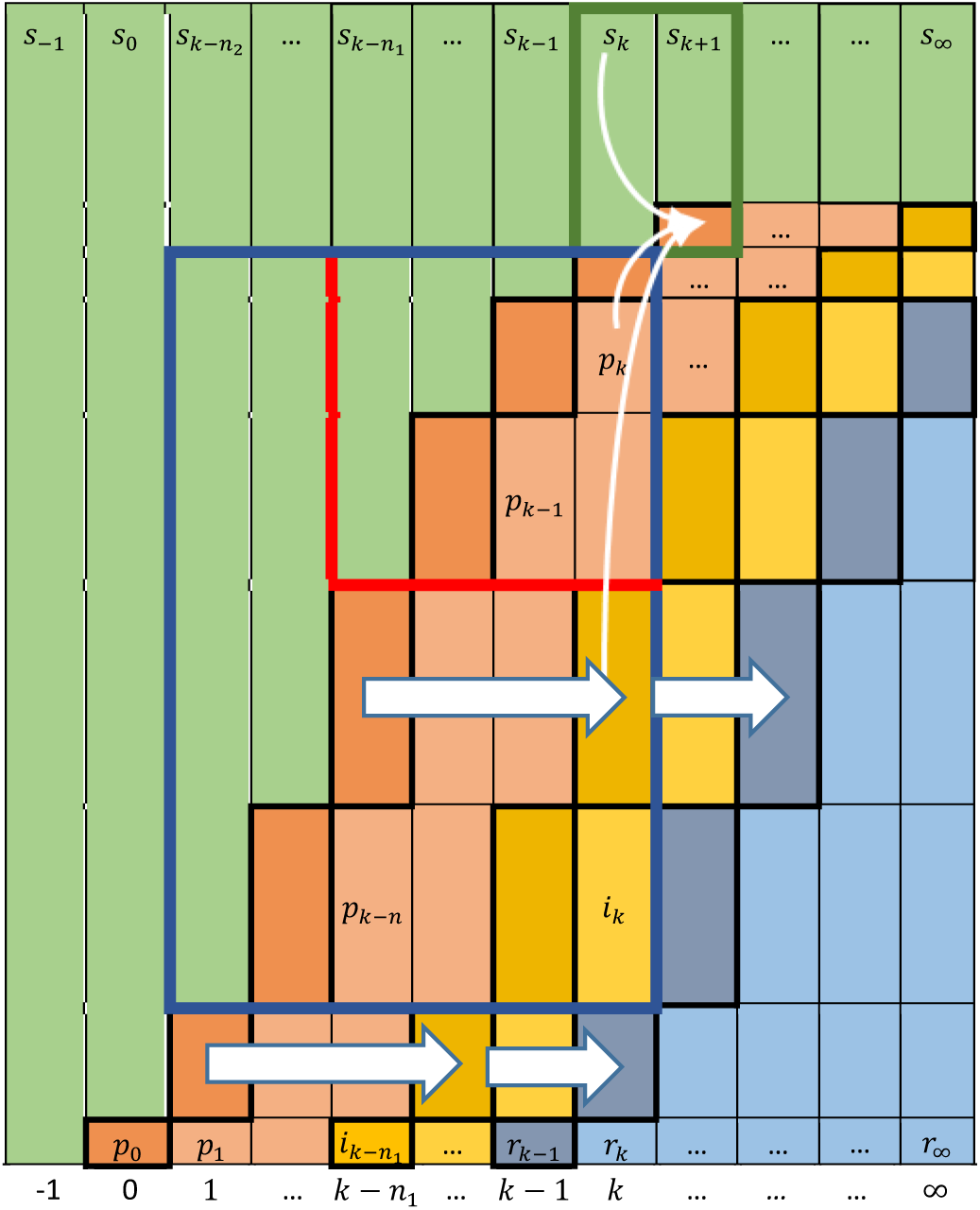
*Schematic of evolving susceptible* (*green*)*, pre-symptomatic infectious* (*red*)*, symptomatic infectious* (*orange*)*, and removed* (*blue*) *fractions of a fixed-size population after an initial infection, p*_0_*. Each new part of a fraction* (*thick-black bordered rectangles*) *moves to the next fraction* (*thick-black bordered rectangles to the right*) *in the same number of time steps. The population eventually reaches a steady state at s_∞_, r_∞_ =* 1 − *s_∞,_ i_∞_ = p_∞_ =* 0.

Note that the form of the SPIR model following the standard SIR pattern, eqns. (1)-(3), is the coupled ODEs^21^

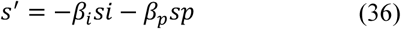

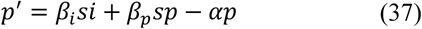

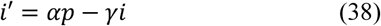

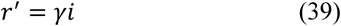

In the form of these ODEs is the following more realistic Padé SPIR model, which approximates the dSPIR DDEs, eqns. (31)-(35), significantly better than the SIR ODEs (Appendix F):

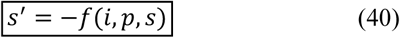

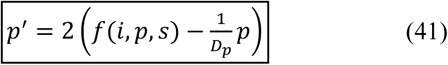

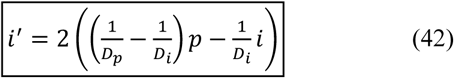

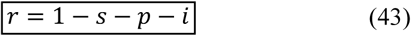

where

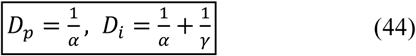

The illustration and analysis presented in sections 3 and 4 can be easily repeated for the dSPIR and Padé SPIR structures. To maintain the scope of this publication, only a few properties will be explored below. The rest will be explored in more detail in forthcoming publications.

### 5.1 From SPIR to dSPIR: Visualization

We present here only a few simulations comparing the {*s, p, i, r*} profiles resulting from numerical solution of the SPIR, dSPIR, and Padé SPIR models. The values

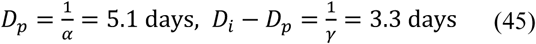

are used in all simulations.^8,9^

Figure 16 illustrates the differences in the infectious peaks, *i^*^* and *p^*^*, similar to these in Figure 8.

**Figure 16.**
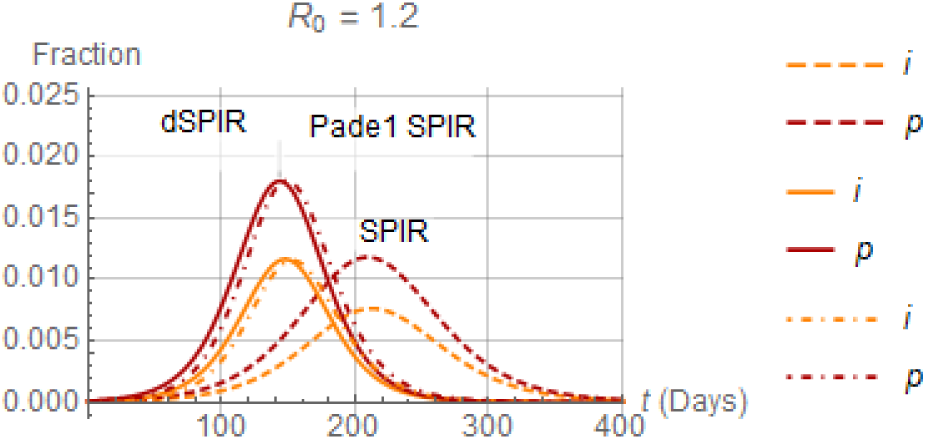
*Comparison of the infectious peaks, i^*^ and p^*^, among the* SPIR (*dashed*), dSPIR (*continuous*) *and* Padé SPIR (*Dot-dashed*) *model structures. Note the proximity between* dSPIR *and* Padé SPIR.

**Figure 17.**
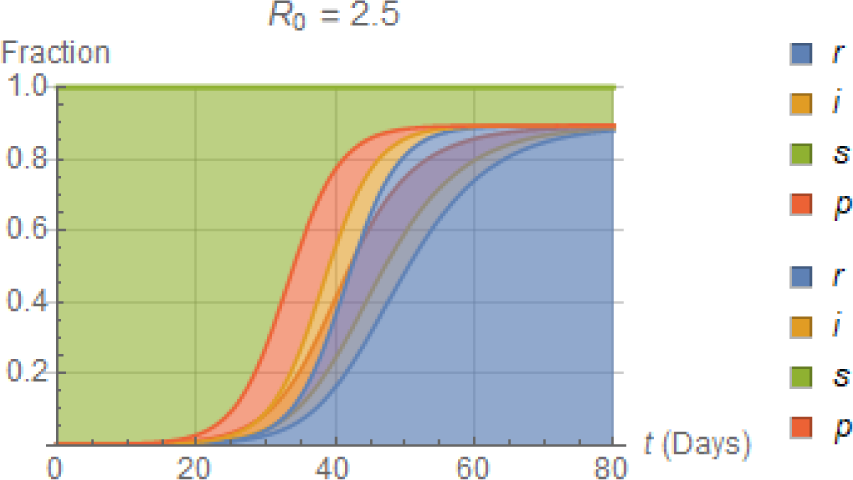
*Comparison of stacked* {*s*, *p, i, r*} *distribution for the* SPIR *and* dSPIR *model structures*.

### 5.2 Stability at equilibrium and epidemic outbreak

Following the same approach as in section 4.1, eqn. (31) can be used to show (Appendix G) that an epidemic outbreak does *not* occur around an equilibrium point 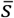 iff

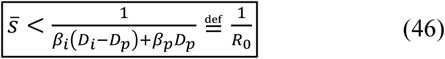

The proof in Appendix G is by approximation. It is conjectured that the bound in the above equation is exact. This conjecture will be examined in subsequent studies.

### 5.3 *Final values of* {*s*, *p, i, r*}

It can be shown (Appendix H) that the dSPIR counterpart of eqn. (19) is

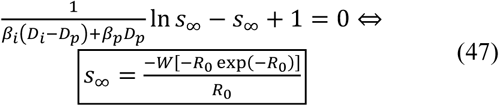

where *D_i_* > *D_p_* and

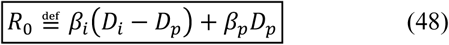

The counterpart of the SIR and dSIR plot (Figure 10) for *r*_∞_ as a function of *R*_0_, eqn. (20), obviously remains the same.

Note that the choice of *β_p_*, *β_i_* has a significant effect on both the stability of the equilibrium point 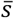, eqn. (46), and on the value of *r_∞_* = 1 − *s_∞_*, eqn. (47), with obvious implications for distancing measures.^21^ For

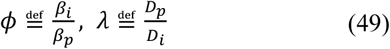

with 0 ≤ *ϕ*, *λ* ≤ 1, eqn. (48) yields

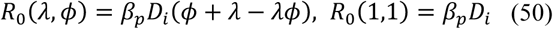

as shown in Figure 18.

**Figure 18.**
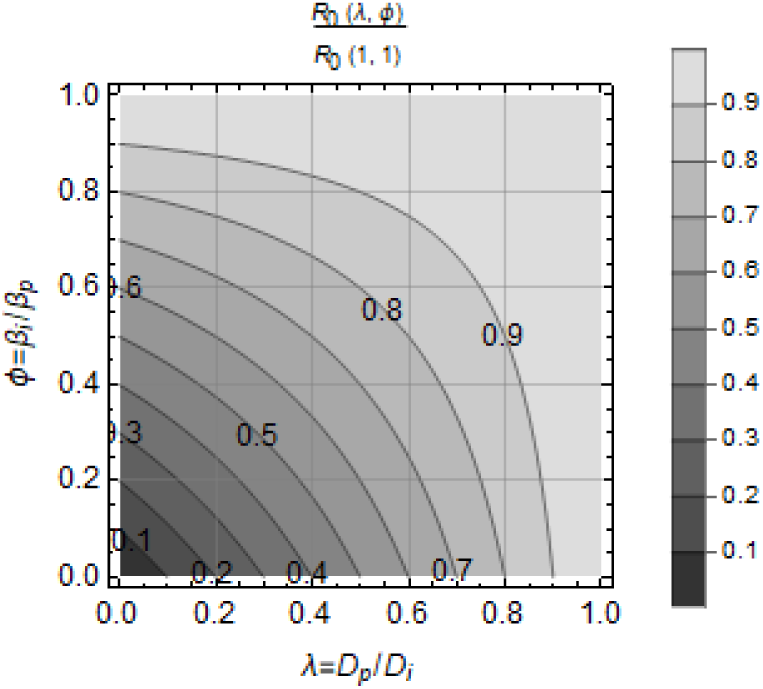
*Contour plot of R*_0_(*λ, ϕ*)/*R*_0_(1,1), *eqn*. (50)*, for the range of additional restrictions*, 0, *placed on symptomatic infectious, and for the possible range of the pre-symptomatic infectious period, D_p_, as a fraction, λ, of the total infectious period, D_i_*.

## 6. Discussion

A subtle inconsistency in the standard SIR model structure was pointed out. This inconsistency arises from misinterpretation of an assumption explicitly articulated in the original publication of Kermack and McKendrick:^1^ that the depletion of the infectious compartment is proportional to the content of that compartment. The depletion rate constant, *γ*, is usually interpreted as equal to the inverse of the residence time in the infectious compartment, namely the duration of the infection, *D*, for each individual (eqn. (18)). To the extent that this duration is about constant, the analysis presented here suggests that the preceding interpretation is incorrect, leading to certain erroneous conclusions.

A corresponding model structure, termed dSIR, was developed, to account for each individual leaving the infectious compartment after a certain duration. The dSIR model structure comprises a single DDE for the susceptible fraction, *s*, and associated algebraic equations capturing the dependence of the remaining population fractions on *s*. While both SIR and dSIR produce the same results for assessment of stability and final values, the SIR model produces a maximum of the infectious fraction, *i^*^*, about half of its dSIR counterpart. This has profound consequences if the SIR model is used to predict *i^*^* during an epidemic. It is also noted that even if the SIR model parameters *β* and *γ* are estimated based on experimental data fit – albeit under the wrong interpretation – model predictions are still going to be inaccurate. This is because the standard SIR model, comprising the three ODEs in eqns. (1)-(3), is structurally different from the DDE form of the dSIR model, eqns. (7)-(9), or even from the ODE form of the Padé-based approximation of the dSIR model DDEs, eqns. (1) and (13) or (1) and (14).

The dSIR structure can be easily extented to other compartment-based population models.^3^ Such an extension to the dSPIR model structure was presented and briefly illustrated and analyzed. This model is important for infections transmitted by both pre-symptomatic and symptomatic infected individuals. Similarities and differences between SPIR and dSPIR models are of the same nature as between SIR and dSIR models.

Numerous additional issues related to this work can be considered, including the following: Rigorous analysis of DDE models; DDE modeling and analysis for a distribution rather than uniform delay (cf. Figure 2); resolution of conjectures presented in the text; and implications for different forms of infection kinetics. Such issues will be addressed in forthcoming publications.

## Data Availability

N/A

## 7. Acknowledgements

All computations were done in *Mathematica*, available at the University of Houston. Sharing of teaching material about the SIR model on Github by Prof. Jeff Kantor of Notre Dame is also gratefully acknowledged. Research reported in this publication was partially supported by the Institute of Allergy and Infectious Diseases of the National Institutes of Health under award number R01AI140287, financed with Federal money. The content is solely the responsibility of the author and does not necessarily represent the official views of the National Institutes of Health.

## Appendix A. Proof of eqns. (13)-(16)

Define the variable

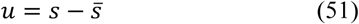

which trivially satisfies *u*(*t*) = 0 for *t* < 0 for simple application of the Translation Theorem of Laplace transforms. Then, take Laplace transforms, 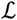, of eqn. (8) with 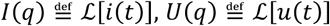, to get

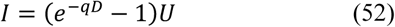

Using the second-order Padé approximation

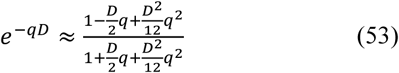

in eqn. (52) yields

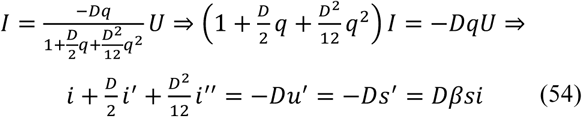

Introducing the new variables 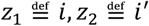 immediately yields eqn. (14).

Eqn. (13) can be proved similarly.

To prove eqn. (16) observe that if *s*(0) changes from its previous value of 1 by a step, −∊, then *U*(*q*) *=* −∊*/q*. Because *r*(*t ≤* 0) = 0 and *s + i + r =* 1, it follows that *i* jumps from *i*(*t <* 0) *=* 0 to *i*(0) *= ∊*. Therefore, the above eqn. (54) yields

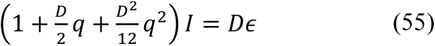

which immediately implies eqn. (16).

Eqn. (15) can be proved similarly.

## Appendix B. Proof of eqn. (17)

Eqns. (7)-(9) imply that (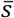, 0, 1, 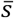) is an equilibrium point, where 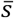 can be arbitrary between 0 and 1. Then, local stability analysis of eqn. (7) by approximate linearization of g(*s_−D_*, *s*) = *f*(*s*, *i*)around the steady state 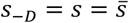 yields

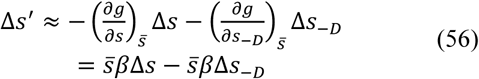

where 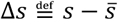 with ∆*s*(*t* < 0) =0, ∆*s*(0) = ∆*s*_0_.

Taking Laplace transforms with 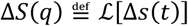 yields 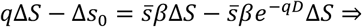

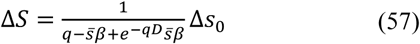

The roots *p_m_* of the transcendental characteristic equation

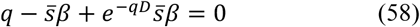

(poles of ∆*S*) can be obtained in terms of the Lambert function,^14,15^ *W*, as follows: The last equation implies

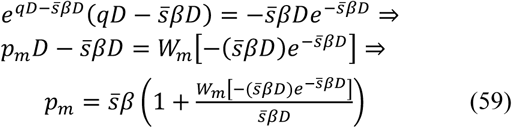

For stability, all *p_m_* must be in the left-half of the complex plane.

We show first that for any positive value of 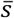, no complex root *p_m_ = λ + jω* can cross the imaginary axis to move from stability to instability. Because, if *p_m_ = jω* were a root of eqn. (58) for 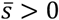, it would be

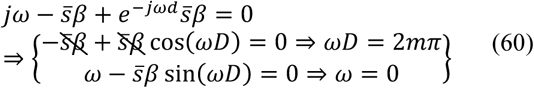

Therefore, only real roots should be considered in eqn. (59) for stability analysis. Furthermore, since

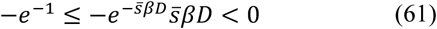

the relevant values of *m* are 0, −1 in eqn. (59) for 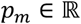, and *p_m_* < 0 implies

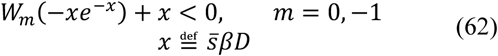

which is satisfied for *x* < 1 (see figure below) leading immediately to eqn. (17).

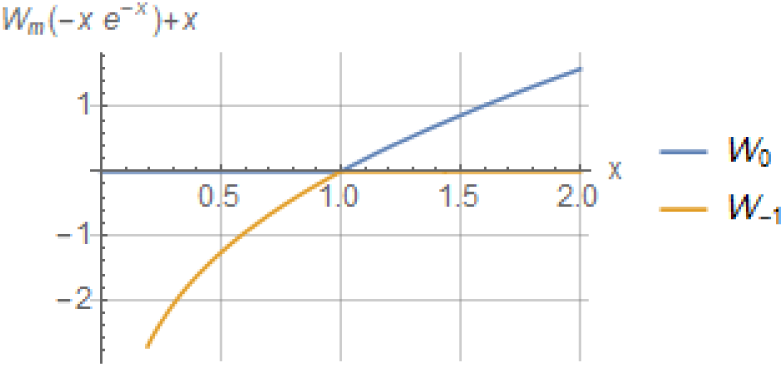

## Appendix C. Proof of eqn. (19)

Dividing eqn. (7) by *s* yields

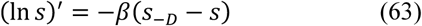

Using eqn. (51) yields *s*_−_*_D_* − *s = u*_−_*_D_* − *u*. Then taking Laplace transforms, 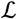, of both sides of eqn. (63) yields

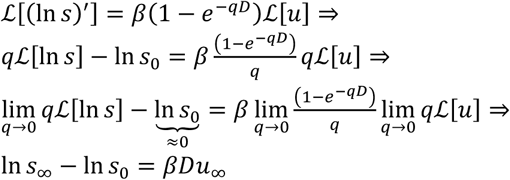

by the Final Value Theorem. Therefore

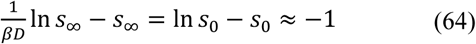

for “small” values of ∊, as shown in the following figure. (Recall that 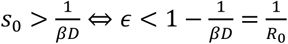 for the epidemic to spread.)

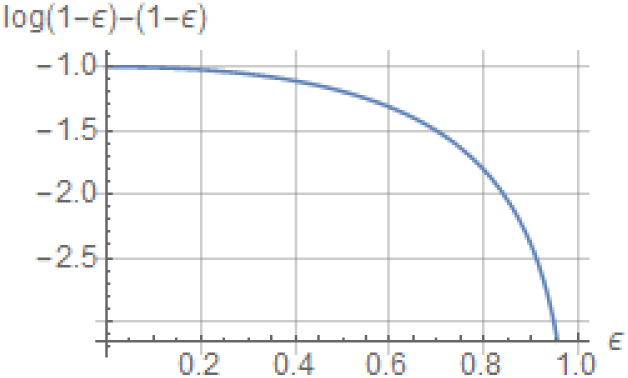

Continuing on the last equation we get

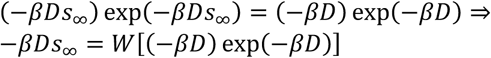

which is eqn. (19).

The Padé SIR model also reaches the same result: Dividing eqn. (1) by eqn. (13) and rearranging yields 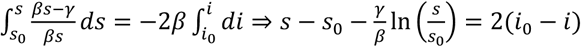

Taking the limit as *t* → ∞ with 1/*γ* = *D*, *s*_0_ ≈ 1, and *i*_0_ ≈ 0 yields eqn. (19).

Proof that the standard SIR model also reaches the same result follows the same pattern and is omitted for brevity.

## Appendix D. Proof of eqns. (21), (22), and (24)

Using the Residue Theorem for Laplace Transforms, eqn. (57) implies

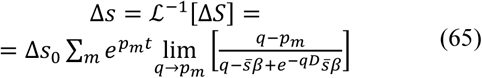

where *p_m_* are the poles of ∆*S*, as shown in eqn. (59).

With 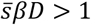 (spreading epidemic) the argument 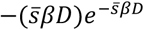 of *W_m_* in eqn. (59) satisfies the inequality in eqn. (61). Consequently, based on the properties of the Lambert function, the summation in eqn. (65) contains two terms with real poles, for *m* = 0, −1, with values

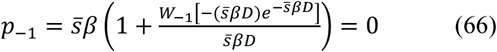

(because *W*_−1_(−*xe*_−_*_x_*) = −*x* for *x* ≥ 1) and

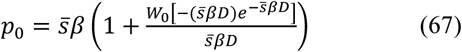

The remaining poles are complex and with negative real parts (see Appendix B) as shown in the following figure of *p_m_ D, m =* −1,0,1,2,…:

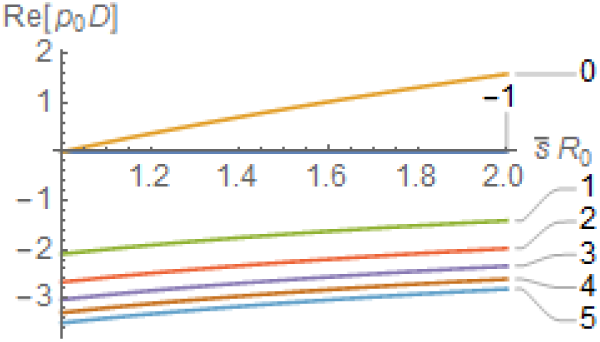

Therefore, the terms *e^pmt^* rapidly decay for *m* ≥ 1, and the summation in eqn. (65) quickly becomes approximately equal to

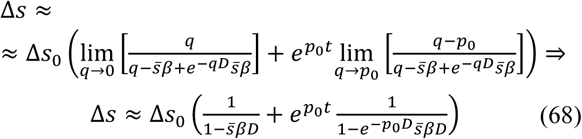

The time dependence of *i* can be obtained in a similar fashion, as eqn. (8), combined with eqn. (57) implies

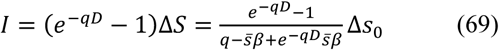

which eventually yields

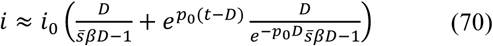

The rates for Padé SIR and standard SIR are obtained by standard ODE linearization analysis and omitted for brevity.

## Appendix E. Proof of eqn.(25)

At the peak value *i^*^* of *i*, eqn. (13) of the first-order Padé SIR model implies

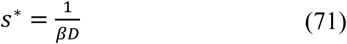

Dividing eqn. (1) by eqn. (13) and integrating yields

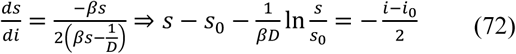

For *i*_0_ = 1 2212 *s*_0_ ≈ 0, combining eqn. (71) with the above yields

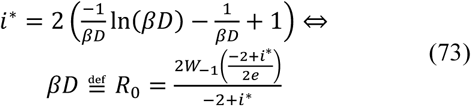

where 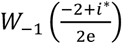 is the Lambert function of order −1, as 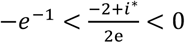 for 0 ≤ *i*^*^ ≤ 1, and 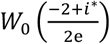 does not yield feasible values above 1.

The peak value *i*^*^ of *i* for the standard SIR model is obtained in an entirely similar way.

## Appendix F. Proof of eqns. (40)-(44)

Following the same approach as in Appendix A for eqn. (32) yields

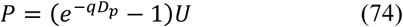

Using the first-order Padé approximation

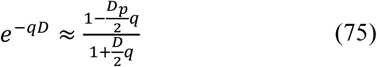

in eqn. (52) yields

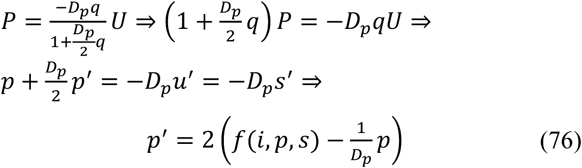

Similarly, adding eqns. (32) and (33), taking Laplace transforms, and using first-order Padé approximation yields

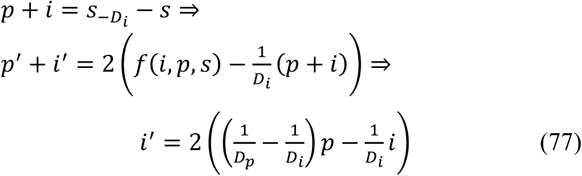

Second-order Padé approximations can be developed in a similar fashion, as shown in Appendix A.

## Appendix G. Proof of eqn. (46)

Eqns. (31)-(34) imply that (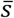, 0, 0, 1 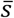) is an equilibrium point, where 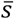 can be arbitrary between 0 and 1. Then, local stability analysis of eqn. (31) by approximate linearization of 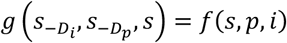 around the steady state 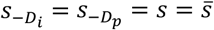 yields

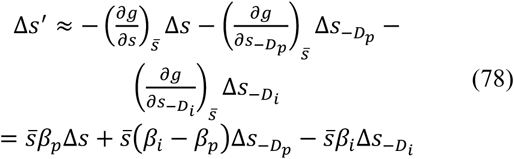

where 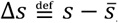 with ∆*s*(*t* < 0) = 0, ∆*s* (0) = ∆*s*_0_.

Taking Laplace transforms with 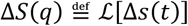 yields

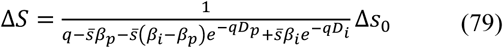

The roots *p_m_* of the transcendental characteristic equation

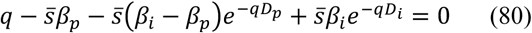

(poles of ∆*S*) cannot be obtained in terms of standard functions known to the author. However, the linear approximation *e^−x^* ≈ 1 − *x* in eqn. (80) immediately yields the result.

## Appendix H. Proof of eqn.(47)

The proof follows the same pattern as in Appendix C, and is presented here briefly.

Dividing eqn. (31) by *s*, using eqn. (51), and taking Laplace transforms yields

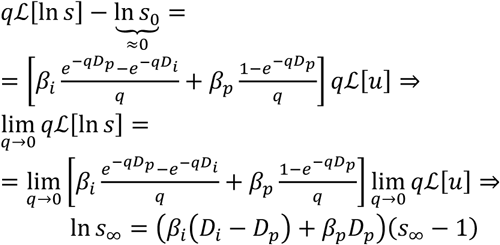

by the Final Value Theorem.

